# Impact evaluation of Lean Healthcare in emergency departments: evidence from a large-scale Brazilian initiative using difference-in-differences

**DOI:** 10.64898/2026.10.16.26365212

**Authors:** Francisco Antônio Sousa de Araújo, Jorge Norio Rezende Ikawa, Alex dos Santos Lima, Livia Muller Bernz, Lucas Reis Correia

## Abstract

**Background:** Lean Healthcare has been adopted as an approach to reducing waste, enhancing process efficiency, and optimizing clinical and administrative workflows. However, empirical evaluations aimed at establishing a causal relationship between Lean Healthcare interventions and outcomes remain limited. This evidence gap is particularly pronounced in the context of large-scale implementations in low- and middle-income countries, where more efficient allocation could yield substantial improvements in population health.

**Methods:** We conducted an impact evaluation study employing a staggered difference-in-differences approach to estimate the effects of the Lean Healthcare in Emergency Departments (ED) initiative in the Brazilian context. The intervention was implemented between January 2017 and December 2025 across 309 hospitals. We analyzed hospital-level indicators of the number of hospitalizations and the average length of stay (in days) among patients admitted through urgent or emergency pathways.

**Results:** Our estimates suggest that the intervention was associated with an average increase of 38.6 admissions per hospital per month (p = 0.033), corresponding to approximately a 5.9% increase relative to the monthly admission volume. We also observed a reduction of 0.184 days in the average length of stay (p = 0.058), equivalent to an approximately 3.0% decrease in hospitalization time. These effects exhibited distinct temporal dynamics: the effect on the number of hospitalizations emerged gradually and increased with the duration of the intervention, whereas the reduction in length of stay became evident earlier.

**Conclusion:** This study provides evidence of a causal effect of Lean Healthcare implementation in emergency departments on operational performance. The simultaneous increase in admissions and reduction in length of stay suggests improved bed productivity and effective capacity without expanding physical infrastructure. These findings support the use of flow-management methodologies as an effective strategy for mitigating emergency department overcrowding while increasing service capacity.

**What is already known on this topic:** Lean Healthcare has been increasingly applied to healthcare service delivery and has been associated with measurable operational improvements in participating hospitals.

**What this study adds:** We employ a quasi-experimental design to assess the causal impact of Lean Healthcare in Emergency Departments (EDs). Our results indicate an increase in hospital admissions and a reduction in the average length of stay, resulting in increased hospital capacity.

**How this study might affect research, practice or policy:** Increased service capacity and reduced length of stay have implications for hospital capacity planning, suggesting that flow-management methodologies may increase effective capacity and reduce the need for costly physical bed expansion.

## 1. Introduction

Emergency Department (ED) overcrowding is a persistent challenge for hospitals[1, 2] and represents a significant threat to the timely and efficient delivery of healthcare. As one of the principal entry points into the hospital[3], inadequate management of the ED can have substantial consequences for patients, healthcare professionals, and the institution as a whole. Among the most immediate consequences of overcrowding are delays in medical assessment and treatment, impaired access to appropriate care, and prolonged inpatient length of stay (IPLOS). Furthermore, ED overcrowding has been associated with increased exposure to procedural errors[4, 5], greater risk of workplace violence against healthcare professionals[6], and potentially higher mortality rates[7].

Strategies proposed to mitigate ED overcrowding range from coordinated interventions involving other healthcare networks and services[8] to the implementation of predictive tools to support patient triage[9, 10]. Some originated in other fields, such as Lean management, which emerged from the Toyota Production System (TPS), developed in post-war Japan and is grounded in the elimination of three fundamental forms of inefficiency: waste (*muda*), unevenness (*mura*), and overburden (*muri*)[11]. Since the early 2000s, Lean principles have been increasingly adapted to healthcare settings, giving rise to the field of Lean Healthcare.

Focusing on the elimination of waste in healthcare processes, the optimization of workflows, and the enhancement of value from the patient’s perspective, Lean Healthcare has been increasingly applied to health service delivery. Systematic reviews[12, 13, 14, 15, 16] have indicated that Lean Healthcare interventions are associated with reductions in IPLOS, thereby supporting their potential application in the redesign and optimization of ED workflows.

Nevertheless, the existing literature has predominantly relied on pre-post intervention comparisons, providing comparatively limited evidence regarding the causal attribution of observed effects to the intervention itself. A notable exception is the study by Vermeulen et al. [17], which compared estimates of ED length of stay obtained from before-after models with those derived from a difference-in-differences approach. The authors reported significantly greater reductions in ED length of stay when using pre-post in comparison to the difference-in-differences approach, highlighting the importance of employing quasi-experimental designs to strengthen causal inference in evaluations of Lean Healthcare interventions.

This study contributes to the existing literature by examining the implementation of Lean principles in ED within the Brazilian healthcare context. It strengthens the evidence base regarding the effects of Lean interventions in EDs by employing a quasi-experimental design, thereby enhancing causal inference and providing a more robust assessment of the intervention’s impact. Specifically, it evaluates a large-scale initiative implemented nationwide through the Institutional Development Support Program of the Unified Health System (Proadi-SUS), a public-private partnership involving private hospitals and the Brazilian Ministry of Health. Under this arrangement, certified philanthropic hospitals are granted federal tax exemptions in exchange for contributing their managerial and technical expertise to strengthen institutions within the Unified Health System (SUS), through projects in areas such as management support, health technology assessment, education, and research.

The primary objective of this study is to evaluate the impact of implementing Lean Healthcare on indicators related to hospital admissions originating from the ED. We hypothesize that optimizing patient flow and organizing the physical workspace in a high-demand ED may generate benefits beyond reductions in IPLOS, including increased service capacity. As a secondary objective, we evaluated whether process optimization through Lean Healthcare was associated with reduced length of stay among hospitalized patients admitted through the ED. This effect may occur through two potential mechanisms: (1) more timely care may allow patients to be treated before their clinical conditions deteriorates, reducing the likelihood of prolonged hospitalization; and (2) standardized protocols managing patients eligible for discharge may promote more efficient and timely transitions from hospital to home. By examining these outcomes jointly, the study also assess whether Lean implementation may increase effective hospital capacity without requiring proportional expansion of physical infrastructure. Thus, our research question is to investigate the impacts of Lean Healthcare on hospital admissions. We hope the findings of this study may contribute to a more comprehensive empirical understanding of the potential indirect effects of Lean Healthcare implementation, particularly within the context of EDs.

## 2. Methods

### 2.1. Data sources

This study relies on two primary data sources: administrative data collected by the project and publicly available data from the Department of Informatics of the Unified Health System (DATASUS). From the project’s administrative records, we extracted the date on which each institution joined the project and its National Registry of Health Facilities (CNES) identifier, a unique identifier assigned to each Brazilian health facility.

Using these identifiers, we collected information on facility characteristics, including location and bed count (both from CNES database), as well as the number and length of stay of urgent/emergency and elective hospitalizations (SIH-RD database). The latter two variables are the project’s outcomes of interest: the number of hospitalizations, which measures the monthly volume of hospital admissions, and the average length of stay, which measures the mean duration, in days, of hospitalizations completed during the month. Both outcomes were aggregated monthly at the hospital level and stratified by type of hospitalization (urgent/emergency and elective).

We thus constructed a panel of 309 hospitals participating in the Lean Healthcare in ED project, covering 108 months from January 2017 through December 2025. We used exclusively secondary data, comprising administrative data aggregated at the hospital unit level and publicly available, anonymized data. No individual-level or personally identifiable information was accessed. Therefore, in accordance with Brazilian Resolution CNS No. 510/2016, which exempts research using exclusively anonymized secondary data from ethical review, this study did not require approval from a Research Ethics Committee. This article follows the guidelines from the Transparent Reporting of Evaluations with Non-randomized Designs (TREND) statement[18].

### 2.2. Project background and intervention

The Lean Healthcare in ED project was adopted by the Ministry of Health in 2017 as an institutional strategy to address overcrowding in EDs of public and philanthropic hospitals. The project operates across four main fronts: (i) detailed diagnosis of patient care workflows; (ii) capacity building and training for multidisciplinary teams; (iii) use of visual management tools and process analysis; and (iv) implementation of rapid improvements (Kaizen events)[19]. These components were intended to improve flow, reduce process variation and delays, and strengthen the ability of multidisciplinary teams to identify and address operational constraints.

The intervention is structured around the DMAIC model (Define, Measure, Analyze, Improve, and Control), which is applied sequentially to the care workflows of participating institutions. The process begins with a preliminary assessment of the institution’s profile, followed by an opening forum during which the project’s objectives, methodology, and results from previous cycles are presented.

Next, a demand and capacity diagnosis is conducted to collect and analyze key clinical and operational data from the hospital. Team training is provided through the High-Performance Management in ED (GAPE program), which incorporates practical simulations contrasting ED settings without structured management strategies with scenarios in which Lean tools have been implemented. Implementation follows a hybrid approach that combines in-person and remote activities conducted by a consulting team comprising physicians and Lean methodology specialists.

In total, 309 hospitals participated in the project between its inception and the end of 2025, covering 205 municipalities across all five regions of Brazil. Hospitals entered the project at different points in time and were organized into successive implementation cycles. **Figure 1** presents the geographic distribution of hospitals participating in the project. We provide a summary of hospitals’ characteristics in the supplementary material.

**Figure 1:**
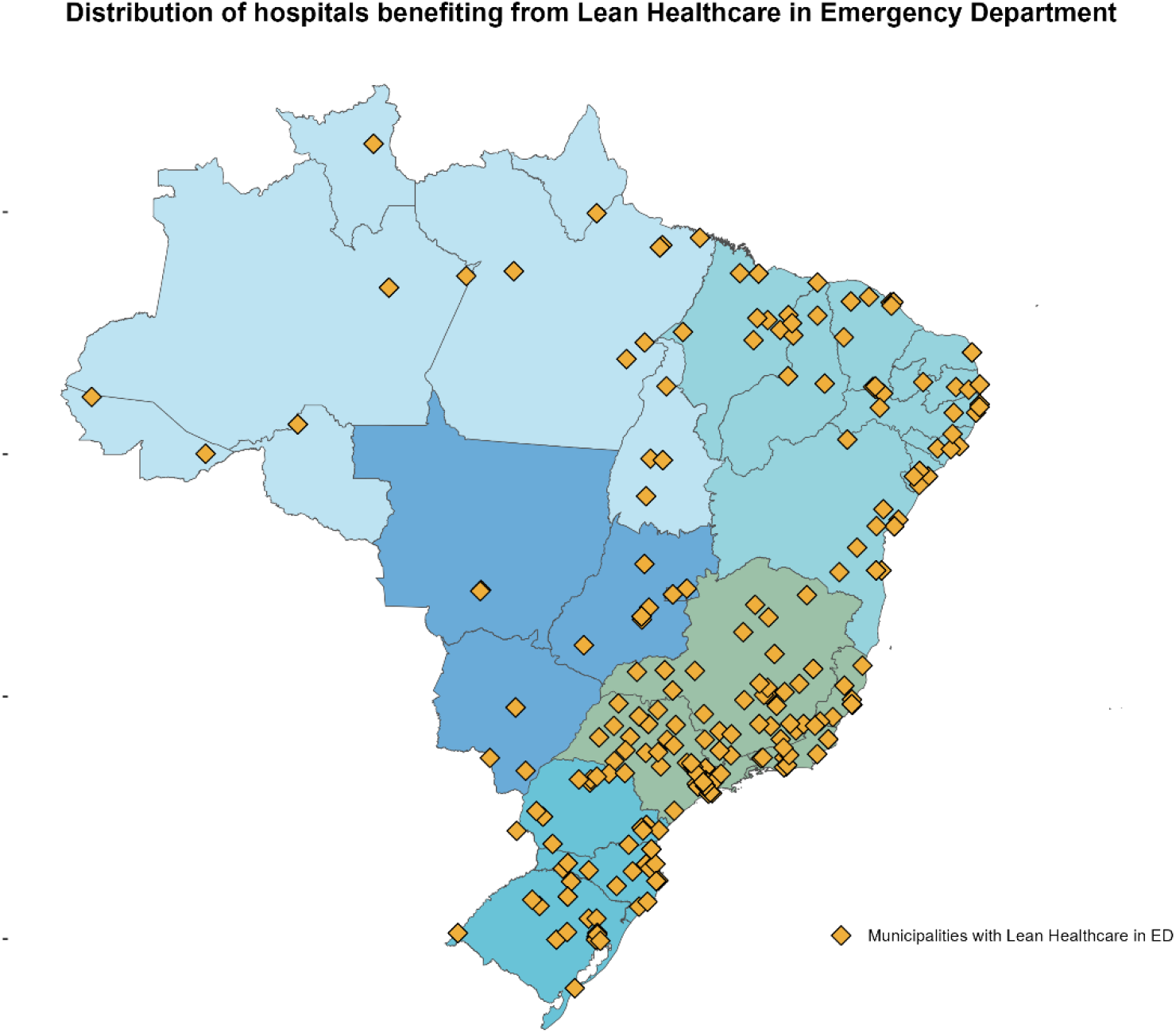
Distribution of hospitals benefiting from Lean Healthcare in Emergency Department.

### 2.3. Statistical methods

We employed a quasi-experimental design to estimate the impact of the project. Exploiting its staggered adoption, we applied the difference-in-differences estimator proposed by Callaway and Sant’Anna[20], using hospitals that had not yet adopted the intervention as the counterfactual group. **Figure 2** presents the cumulative distribution of hospitals treated by the project over time. The difference between consecutive bars represents hospitals newly entering the project in each operational cycle.

**Figure 2:**
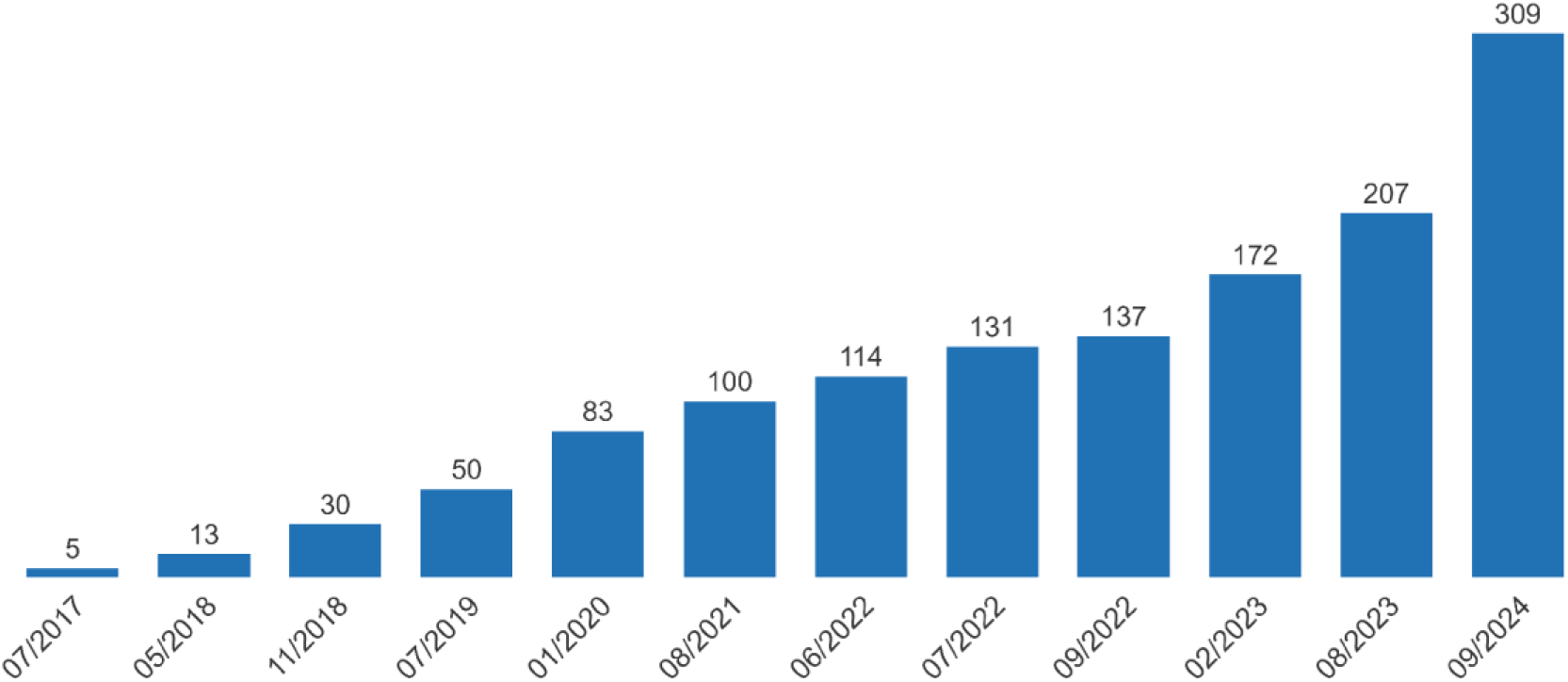
Cumulative distribution of hospitals treated by the project in each operational cycle.

Following Callaway and Sant’Anna approach, let *i* index hospitals and *t* index months in the panel, while *G*_*i*_ denotes the month in which hospital *i* joined the Lean Healthcare in ED project. The potential outcome *Y*_*it*_ (*g*) represents the outcome for hospital *i* in month *t* if it entered the project in month *g*, whereas *Y*_*it*_ (0) denotes the potential outcome in the absence of treatment throughout the observation period. The parameter of interest is the average treatment effect on the treated for cohort *g* in period *t*:

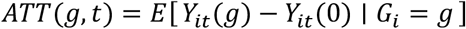

Identification of this parameter relies on three assumptions. First, the absence of anticipation requires that hospitals do not alter their outcomes before the month in which they enter the project. Second, the parallel trends assumption requires the potential outcomes of cohort *g* and the comparison group to follow parallel trajectories in the absence of treatment, allowing the counterfactual path to be extrapolated. Third, the common support assumption requires the existence of comparable untreated units for each cohort-period cell. Under these assumptions, the parameter is identified as the difference between the changes in outcomes for the treated and comparison groups, with both changes measured relative to the last month before cohort *g*’s entry:

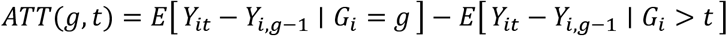

The main specification does not include covariates. In this setting, the estimator is given by the difference between the mean changes in outcomes for the treated and comparison groups.

The panel contains no never-treated hospitals. All 309 hospitals received intervention at some point between January 2017 and December 2025. Consequently, the composition of the final cohort directly affects the sample used to identify the treatment effect. The 102 hospitals that entered the project in September 2024 (33.0% of the panel) served as the comparison group for all preceding cohorts, but do not contribute to post-treatment estimates because no untreated hospitals remain after their entry. August 2024 is therefore the last month for which a comparison group is available, and treatment effects are identified using the 207 hospitals in the first eleven cohorts.

## 3. Results

### 3.1. Descriptive analysis of outcome variables

Table 1. reports the mean and standard deviation (in parentheses) of the outcome variables, number of hospitalizations and average length of stay (in days) for patients admitted through urgent or emergency pathways, aggregated at the monthly level for the 309 participating hospitals over the 108-month study period, by treatment period (pre-and post-treatment), and by geographic region. On average, participating hospitals recorded 751 hospitalizations among patients admitted through urgent or emergency pathways, with an average length of stay of 6.26 days. The disaggregated results reveal substantial regional variation in the monthly volume of such hospitalizations.

**Table 1:** Outcome variables of hospitals participating in the Lean Healthcare in ED.

| **Primary outcome (urgent or emergency)** | **All period** | **Not yet treated period** | **Treated period** |
| --- | --- | --- | --- |
| Number of Hospitalizations | 751<br>(468.33) | 654.88<br>(412.41) | 902.08<br>(509.33) |
| Average length of stay (days) | 6.26<br>(2.21) | 6.15<br>(2.3) | 6.43<br>(2.04) |
| Number of observations | 33,372 | 20,399 | 12,973 |
| **Primary outcome (urgent or emergency) by region** |  |  |  |
| **Central-West (23 hospitals - 7.44% from the sample)** |  |  |  |
| Number of Hospitalizations | 968.89<br>(598.55) | 758.15<br>(490.84) | 1195.11<br>(620.98) |
| Average length of stay (days) | 6.03<br>(3.18) | 6.08<br>(4.14) | 5.97<br>(1.61) |
| Number of observations | 2,484 | 1,319 | 1,165 |
| **North (26 hospitals - 8.41% from the sample)** |  |  |  |
| Number of Hospitalizations | 581.08<br>(319.18) | 515.16<br>(256.54) | 670.92<br>(370.29) |
| Average length of stay (days) | 6.8<br>(2.63) | 6.69<br>(2.58) | 6.96<br>(2.7) |
| Number of observations | 2,808 | 1,612 | 1,196 |
| **Northeast (81 hospitals - 26.21% from the sample)** |  |  |  |
| Number of Hospitalizations | 740.77<br>(550.68) | 646.67<br>(506.53) | 887.06<br>(583.71) |
| Average length of stay (days) | 6.57<br>(2.35) | 6.32<br>(2.48) | 6.96<br>(2.07) |
| Number of observations | 8,748 | 5,339 | 3,409 |
| **South (63 hospitals - 20.39% from the sample)** |  |  |  |
| Number of Hospitalizations | 740.59<br>(385.56) | 638.54<br>(310.83) | 894.17<br>(433.32) |
| Average length of stay (days) | 5.86<br>(1.64) | 5.85<br>(1.6) | 5.87<br>(1.7) |
| Number of observations | 6,804 | 4,045 | 2,759 |
| **Southeast (116 hospitals - 37.54% from the sample)** |  |  |  |
| Number of Hospitalizations | 759.86<br>(427.66) | 680.54<br>(392.21) | 903.5<br>(451.14) |
| Average length of stay (days) | 6.19<br>(1.98) | 6.1<br>(1.97) | 6.35<br>(1.98) |
| Number of observations | 12,528 | 8,084 | 4,444 |

Standard deviation in parentheses.

### 3.2. Treatment effect

**Table 2** reports the estimated effects on the number of hospitalizations and average length of stay (days), separately for patients admitted through urgent or emergency pathways (“urgent type”) and elective admissions (“elective type”). Given that the intervention was implemented in the ED, we hypothesized that its effects would be limited to urgent or emergency admissions. Elective admissions, which were not exposed to the intervention, served as a placebo test, for which no statistically significant effect was expected.

**Table 2:** Estimated treatment effect.

| Outcome | Type | ATT | Standard<br>-error | CI 95% | p-value | Mean<br>before | Relative<br>effect | Observ. |
| --- | --- | --- | --- | --- | --- | --- | --- | --- |
| Number of<br>hospitalizations | Urgent | 38.608* | 18.129 | [3.074; 74.141] | 0.033 | 654.88 | 5.9% | 33,005 |
|  | Elective | -6.696 | 8.239 | [-22.843; 9.452] | 0.416 | 163.87 | -4.1% | 28,472 |
| Average length<br>of stay (days) | Urgent | -0.184* | 0.097 | [-0.374; 0.006] | 0.058 | 6.15 | -3.0% | 33,005 |
|  | Elective | -0.245 | 0.221 | [-0.678; 0.188] | 0.267 | 3.10 | -7.9% | 28,472 |

\* $p < 0,10$ .

We estimate that the project increased the monthly volume of urgent or emergency admissions by an average of 38.6 admissions per hospital (p = 0.033; 95% CI: 3.1 – 74.1). Treated hospitals recorded an average of 654.9 emergency admissions per month before joining the project, indicating that the estimated effect corresponds to a 5.9% increase relative to the baseline level. We also find suggestive evidence that the project is associated with a reduction in the average length of stay among urgent or emergency admissions. The estimated effect is a decrease of 0.184 days per admission (p = 0.058; 95% CI: −0.374 – 0.006), equivalent to approximately 4.4 fewer hours per admitted patient. Relative to the pre-treatment baseline of 6.15 days, this estimate represents a reduction of approximately 3.0% in average length of stay. As expected, we find no statistically significant effect among elective admissions, which constitute the non-intervention group.

Figure 3. present the results disaggregated by time, considering the number of hospitalizations (**Panel A**) and average length of stay (in days) (**Panel B**). The estimates are reported as coefficients by month of exposure, using a window spanning the twelve months preceding and following entry, with 95% confidence intervals. Both panels provide evidence of parallel pre-treatment trends, as the pre-entry coefficients are not statistically distinguishable from zero for either outcome.

**Figure 3:**
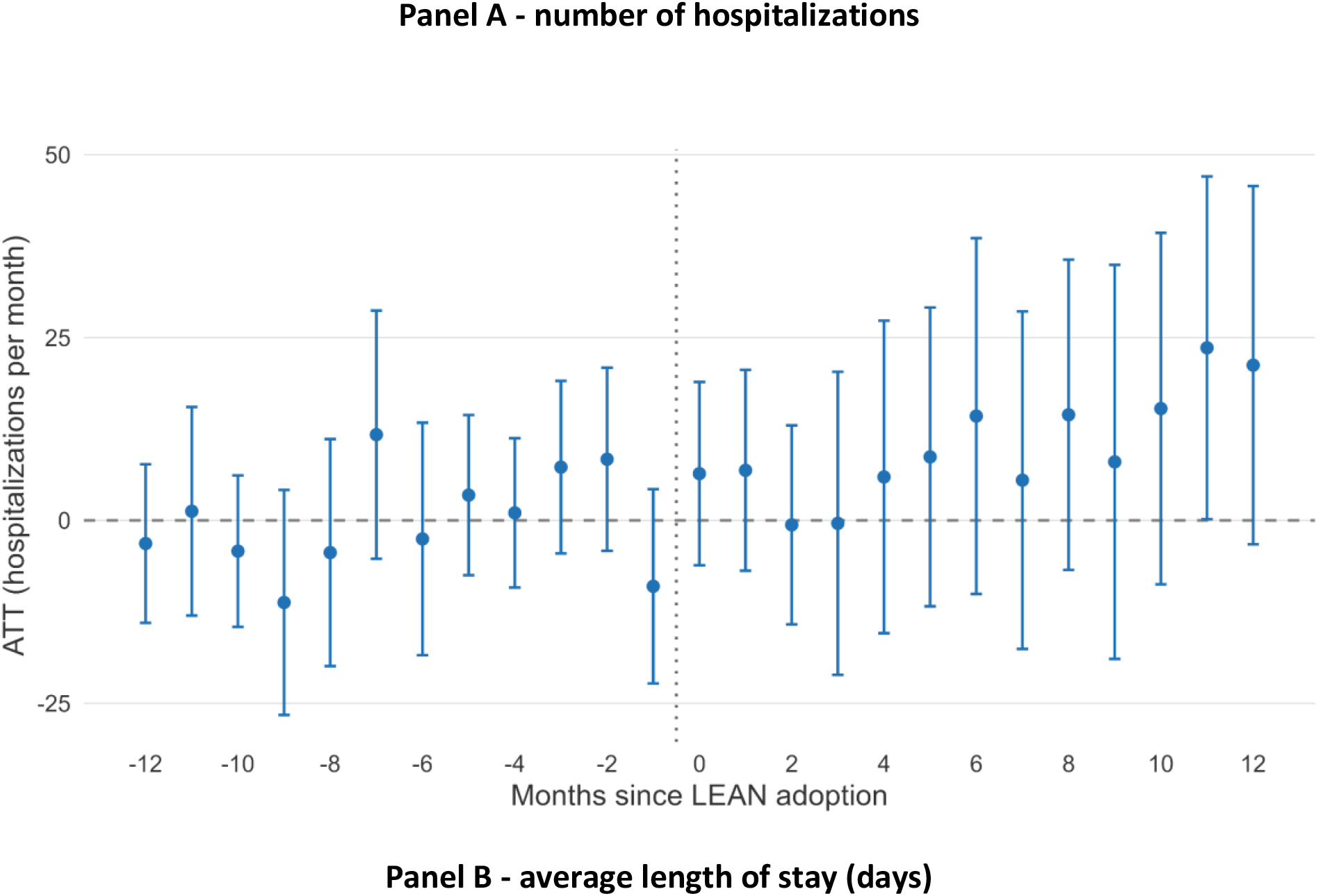
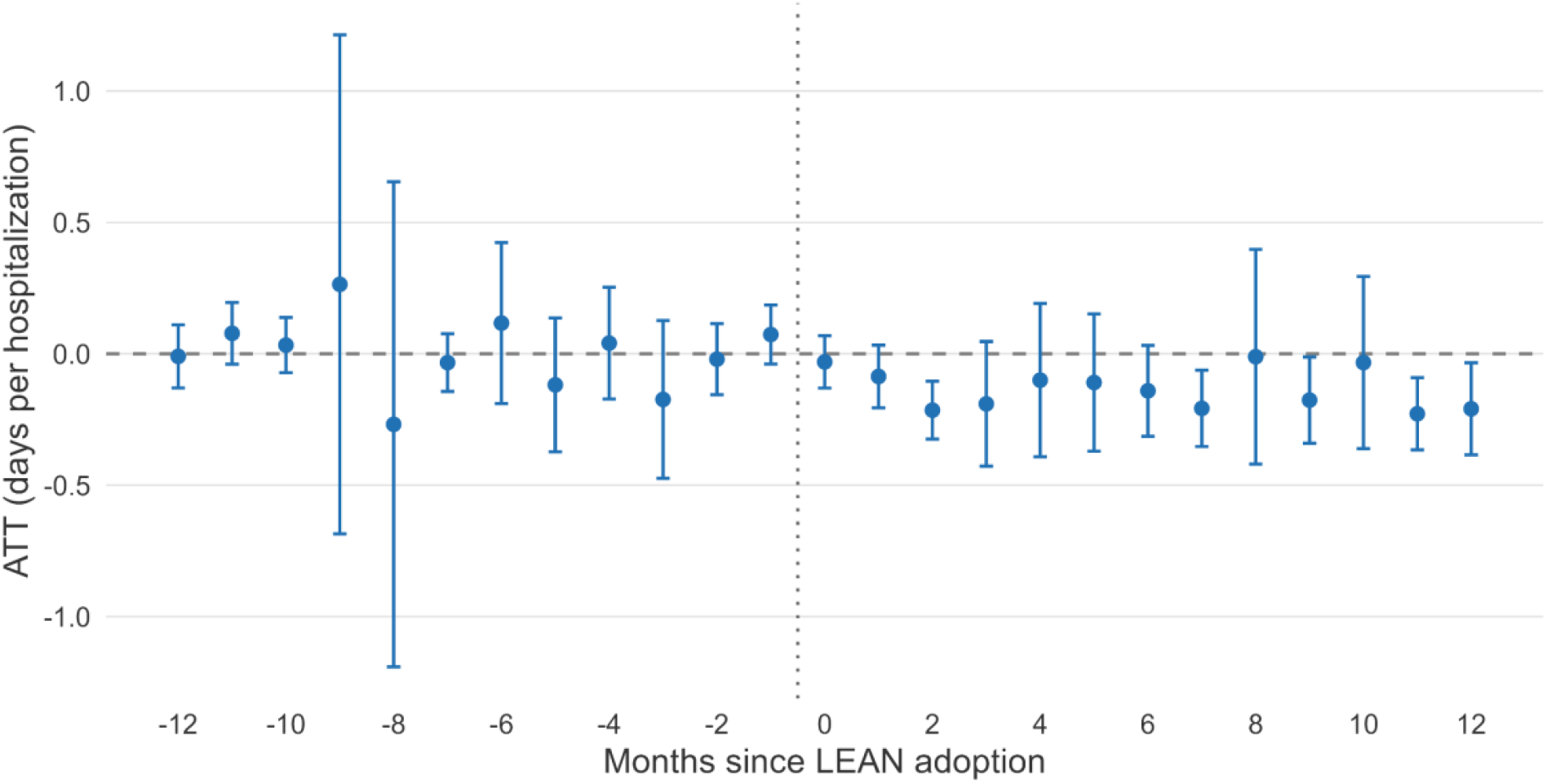
Event study for the number of hospitalizations and the average length of stay (days).

The effect on the number of hospitalizations, reported in **Figures 3 - Panel A**, emerges gradually and increases with the duration of exposure. Coefficients during the initial months remain close to zero, with statistically significant effects appearing only at the eleven- and twelve-month horizons, corresponding to increases of 23.6 and 21.2 hospitalizations, respectively. The dynamic aggregation over the twelve-month window yields an estimated increase of 9.9 hospitalizations (p = 0.206), substantially lower than the 38.6 hospitalizations estimated using the simple aggregation.

The effect on length of stay, reported in **Figures 3 - Panel B**, emerges early and persists throughout the post-treatment period. The coefficient remains negative across all twelve months of the post-treatment window, reaching statistical significance as early as the second month of exposure, with a median effect of −0.14 days over the window. The dynamic aggregation over the twelve-month post-treatment period yields an estimated reduction of 0.134 days (p = 0.078), close to simple aggregation estimate reported in **Table 2**.

The difference between the two aggregation methods reflects different time horizons rather than an inconsistency between the estimates. The simple aggregation **(Table 2)** incorporates all post-treatment months available for each cohort, whereas the event-study is restricted to the first twelve months following treatment. When the effect increases with the duration of exposure, the simple aggregation naturally produces a larger estimates.

The supplementary material reports two sensitivity analyses. The first excludes the initial cohort of five hospitals that participated during the pilot phase. The second restricts the analysis to the subsample of units that received the intervention from June 2022 onward, thereby excluding from the estimation the period characterized by the suspension of elective procedures and overcrowding of emergency departments during the COVID-19 pandemic. Overall, the findings from these sensitivity analyses are consistent with those of the main analysis, although statistical significance is not retained for some estimates, likely reflecting the reduced sample size.

## 4. Discussion

Using a quasi-experimental design, this impact evaluation provides empirical evidence that implementing Lean Healthcare in EDs within the Brazilian context was associated with measurable operational improvements among participating hospitals: an increase in monthly patient volume and a reduction in length of stay among admissions through urgent or emergency pathways. These findings are consistent with previous research[12, 13, 14, 15, 16], predominantly based on pre-post designs, which has associated Lean Healthcare with improvements across several operational dimensions, including workflow efficiency, waiting times, and patient throughput.

Unlike most previous studies, with the notable exception of Vermeulen et al.[17], this study uses hospitals that had not yet received the intervention as a counterfactual, enabling a more rigorous assessment of its effect. In conventional pre-post studies, post-intervention changes may reflect not only the intervention itself but also organizational learning, accumulated experience, or other temporal factors. The difference-in-differences approach accounts for such temporal effects under the assumption that, in the absence of the intervention, outcomes in the treated and control groups would have evolved similarly. It therefore provides a more robust basis for distinguishing intervention-related effects from improvements attributable to the passage of time, yielding estimates that more closely approximate the intervention’s causal effect.

The study design also enabled a month-by-month assessment of effects throughout the year following implementation, revealing distinct temporal patterns across the outcomes examined. Improvements in average length of stay emerged early, as soon as two months after the intervention commenced, whereas effects on hospitalization volume developed gradually and became increasingly pronounced towards the end of the observation period.

This temporal pattern is broadly consistent with the literature on the maturation and sustainability of Lean implementation in healthcare. Studies reporting partial or complete erosion of initial gains over time have attributed such reversals to insufficient integration of Lean principles into organizational strategy and culture[21]. Similarly, reviews of barriers to Lean adoption in developing countries have identified cultural resistance, limited support from senior management, and inadequate formal training as recurrent obstacles[22]. Against this backdrop, the progressive increase in the effect on hospitalization volume through the eleventh and twelfth months of exposure suggests that improvements in patient throughput depend not merely on the isolated adoption of specific Lean tools, but also on a broader process of organizational maturation and the progressive integration of these practices into routine operations. The gradual emergence and strengthening of this effect are therefore consistent with the notion that sustained operational improvements require time for new practices to become embedded within organizational routines, processes, and culture.

Beyond its academic contribution, the finding that effects mature at different rates across outcomes also has important managerial implications. The distinct trajectories observed for average length of stay and hospitalization volume suggest that Lean interventions should be evaluated using outcome-specific time horizons. Because effects on hospitalization volume take longer to mature, assessing a newly implemented initiative on the basis of admission volumes during its first months may underestimate its eventual impact. Such premature assessments could lead decision-makers to discontinue or reduce investment in an intervention before its benefits have fully materialized.

From a management perspective, the combination of increased service capacity and reduced length of stay is particularly relevant to hospital capacity planning. An increase of 5.9% in the number of emergency admissions, combined with a 3.0% reduction in length of stay, indicates an improvement in bed productivity without requiring a corresponding expansion of physical capacity. This finding strengthens the rationale for investing in flow-management methodologies as a means of increasing effective capacity, potentially providing an alternative to the considerably more costly strategy of expanding the physical bed base.

Finally, the absence of an effect on elective admissions, which do not depend on the ED flow targeted by the intervention, offers a form of internal validation, suggesting that the estimated improvements were specific to the patient flow affected by the intervention rather than reflecting broader administrative or operational changes across the hospitals. From a managerial standpoint, this specificity may also strengthen decision-makers’ ability to communicate and justify the intervention’s impact to clinical stakeholders and funding bodies.

Some limitations should be considered when interpreting the findings of this study. The absence of hospitals that remained untreated throughout the panel period is the most important structural limitation of the design. Because the 309 participating hospitals adopted the intervention at different points between 2017 and 2025, identification relies on comparisons between hospitals already exposed and those not yet exposed. The estimates should therefore be interpreted in light of this staggered adoption structure and the assumptions required for valid causal inference.

Second, the causal estimates are restricted to two outcomes: the number of hospitalizations and the average length of stay among patients admitted through urgent or emergency pathways. Other relevant dimensions of hospital performance, such as waiting times, mortality, intensive care unit (ICU) utilization, patient-days, and bed turnover, were not examined. The absence of these outcomes limits the extent to which the study can assess the broader effects of the intervention on hospital performance and patient care.

Third, although the various specifications consistently indicate an increase in hospitalization volume and a reduction in length of stay, statistical significance at the 5% level is not maintained across all models. In the main specification, the effect on average length of stay is significant only at the 10% level (p = 0.058). Similarly, in the event-study analyses with dynamic aggregation, effects on hospitalization volume (p = 0.206) and length of stay (p = 0.078) do not reach conventional significance at the 5% level. These results warrant caution in interpreting the magnitude and precision of the estimated effects.

Fourth, interpreting the increase in hospitalization volume as an improvement in care throughput, rather than greater clinical severity, is plausible and consistent with the simultaneous reduction in length of stay, but the available data do not permit this interpretation to be established conclusively. Finally, participation in the project is preceded by a preliminary assessment of hospitals against minimum criteria related to overcrowding, data availability, and institutional commitment. The estimated effects therefore apply primarily to hospitals meeting these institutional readiness criteria, and caution is warranted when generalizing the findings. This selection process may limit external validity, as the effects observed may not be directly transferable to healthcare facilities with substantially different organizational characteristics or lacking the conditions required for implementation.

This study reinforces existing evidence that Lean methodologies can enhance hospital efficiency by increasing admissions and reducing length of stay. Impact evaluations provide important evidence on causality, however, they represent only one dimension of assessment. Further research using economic, equity, and implementation perspectives can bring a complementary understanding of the intervention’s effects.

## 5. Conclusion

Overall, this study provides evidence of a causal effect, within the Brazilian healthcare context, of introducing Lean Healthcare in emergency departments on the operational performance of participating hospitals. It thereby helps address an important gap in the Lean Healthcare literature, in which intervention effects have more commonly been evaluated using pre-post comparisons rather than quasi-experimental approaches that allow more rigorous identification of causal effects. This impact evaluation thus strengthens the evidence regarding the extent to which the observed operational improvements can be attributed to the intervention itself rather than to concurrent temporal trends or other factors.

The study also contributes to a more nuanced understanding of the temporal dynamics of Lean implementation: the effects of Lean Healthcare in emergency departments do not necessarily emerge or mature at the same rate across outcomes. This underscores the importance of adopting outcome-specific time horizons when evaluating the effectiveness and sustainability of healthcare interventions. More broadly, the findings suggest that assessments conducted too early in the implementation process may fail to capture the full magnitude of the benefits of Lean interventions, particularly for outcomes whose effects take longer to mature.

## Data Availability

All data produced in the present study are available upon reasonable request to the authors

## Funding and Conflicts of interest

The resources for conducting this research were provided by Proadi-SUS (Programa de Apoio ao Desenvolvimento Institucional do Sistema Único de Saúde). Therefore, the authors manifest an indirect institutional conflict of interest as they are affiliated to hospitals in partnership with the Brazilian Ministry of Health for this program and developing the Lean Healthcare Project as well as its evaluation.

Since the data was publicly available no ethical committee approval was needed. Furthermore, the protocol for the impact evaluation was registered in https://ridie.3ieimpact.org/index.php.

## Contributions

Francisco Antônio Sousa de Araújo: Conceptualization, methodology, investigation, formal analysis, data curation, visualization, and review.

Jorge Norio Rezende Ikawa: Conceptualization, methodology, investigation, formal analysis, visualization, writing original draft, and review.

Alex dos Santos Lima: Methodology, data curation and formal analysis. Livia Muller Bernz: Investigation, interpretation of findings and review. Lucas Reis Correia: Conceptualization, methodology, writing and review.

## Acronyms

ED: Emergency Departments

## Supplementary material

**Table 1A** presents the mean values for participating hospitals, with standard deviations in parentheses. Municipal population data are derived from the 2022 Brazilian Census. “Number of hospitalizations” represents the monthly average number of hospitalizations from January 2017 to December 2025. The remaining data are sourced from CNES, with January 2026 as the reference date.

**Table 1A:** Descriptive analysis of hospitals participating in the Lean Healthcare in Emergency Department.

|  | All sample | Central-West | North | Northeast | South | Southeast |
| --- | --- | --- | --- | --- | --- | --- |
| Number of hospitals | 309 | 23 | 26 | 81 | 63 | 116 |
| Municipal population | 1,188,491<br>(2,384,584) | 1,313,151<br>(1,111,770) | 732,643<br>(751,039) | 798,008<br>(880,668) | 484,549<br>(539,081) | 1,920,924<br>(3,633,714) |
| Number of hospitalizations | 898.56<br>(560.94) | 1053.51<br>(592.47) | 636.78<br>(281.44) | 832.97<br>(602.66) | 942.07<br>(490.46) | 948.68<br>(590.86) |
| Number of beds | 264.78<br>(178.33) | 309.78<br>(198.01) | 198.04<br>(94.68) | 255.30<br>(185.26) | 268.10<br>(123.10) | 275.64<br>(205.13) |
| Number of beds for the SUS | 237.25<br>(168.84) | 293.70<br>(189.89) | 186.88<br>(98.39) | 246.72<br>(185.17) | 219.84<br>(111.03) | 240.20<br>(188.15) |
| Number of ICUs | 40.94<br>(29.24) | 41.70<br>(29.97) | 27.69<br>(19.34) | 35.49<br>(28.41) | 42.67<br>(21.83) | 46.63<br>(33.52) |
| Number of ICUs for the SUS | 34.70<br>(26.88) | 36.83<br>(28.97) | 21.85<br>(18.09) | 31.72<br>(26.57) | 34.24<br>(17.58) | 39.50<br>(31.27) |

**Table 2A** presents the results of the sensitivity analyses, including a specification that excludes the first cohort of hospitals (i.e., the “without initial cohort” specification) and a specification restricted to units treated from June 2022 onward (i.e., the “starting in June 2022” specification). For comparison, the table also reports the results from the main specification. Overall, the results of the sensitivity analyses are consistent in direction with those of the main specification, although statistical significance is not maintained across all estimates.

**Table 2A:**
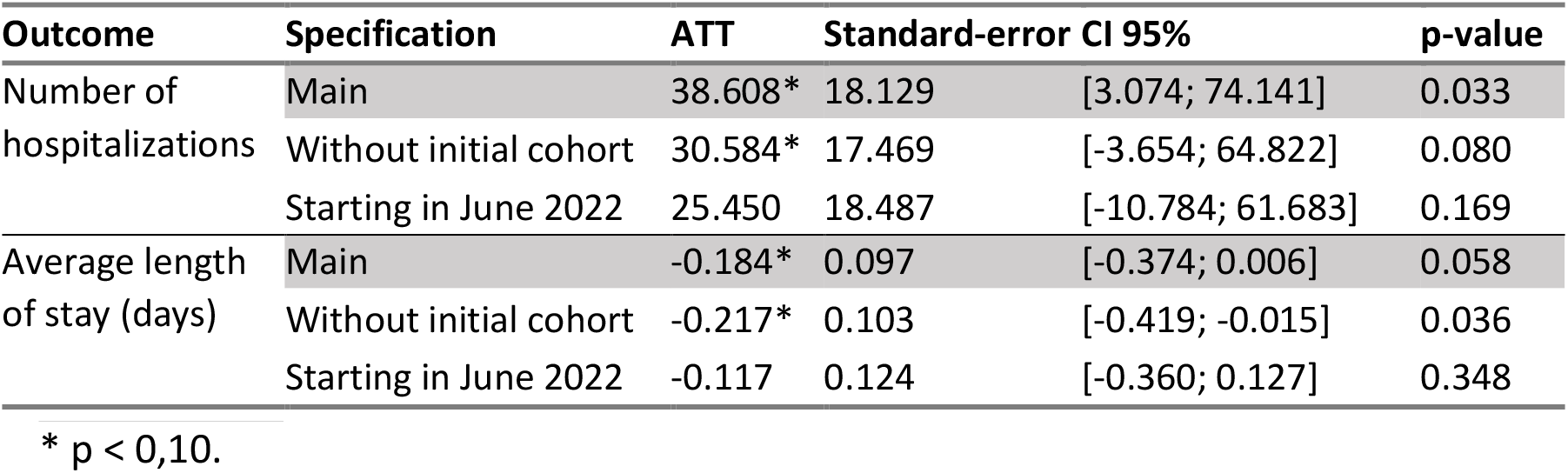
Sensitivity analyses of estimated treatment effect.

Excluding the first cohort leaves both results with similar magnitude. When hospitals that joined the project in July 2017 are excluded, the estimated effect on the volume of emergency admissions is 30.6 (p = 0.080), while the estimated effect on average length of stay is −0.217 days (p = 0.036). These results indicate that the oldest cohort, which has the longest exposure horizon in the panel, does not drive either of the main findings.

The post-pandemic subsample preserves the direction of both effects but yields smaller estimated magnitudes. Restricting the sample to observations from June 2022 onward produces an estimated effect of 25.4 (p = 0.169) on the volume of emergency admissions and −0.117 days (p = 0.348) on average length of stay. Although both estimates retain the signs observed in the main specification, neither is statistically significant, likely reflecting the reduced sample size.

The loss of statistical significance arises through different channels for the two outcomes. For emergency admission volume, the estimated coefficient decreases by 34%, while the standard error remains virtually unchanged relative to the main specification, differing by only 2%. For average length of stay, the estimated coefficient decreases by 37%, while the standard error increases by 28%. The post-pandemic subsample excludes cohorts that joined the project before June 2022 and consequently shortens the exposure horizon for the remaining cohorts. As shown in the results section, the estimated effect on admission volume is concentrated precisely at longer exposure horizons.

## References

1. Morley C, Unwin M, Peterson GM, et al. Emergency department crowding: a systematic review of causes, consequences and solutions. PLoS One. 2018;13(8):e0203316. doi:10.1371/journal.pone.0203316.

2. Sartini M, Carbone A, Demartini A, et al. Overcrowding in emergency department: causes, consequences, and solutions—a narrative review. Healthcare (Basel). 2022;10(9):1625. doi:10.3390/healthcare10091625.

3. Studnicki J, Platonova EA, Fisher JW. Hospital-level variation in the percentage of admissions originating in the emergency department. Am J Emerg Med. 2012;30(8):1441–6. doi:10.1016/j.ajem.2011.11.008.

4. Lee CC, Lee MG, Hsu TC, et al. The impact of overcrowding on the bacterial contamination of blood cultures in the ED. Am J Emerg Med. 2012;30(6):839–45. doi:10.1016/j.ajem.2011.05.026.

5. Kulstad EB, Sikka R, Sweis R, et al. ED overcrowding is associated with an increased frequency of medication errors. Am J Emerg Med. 2010;28(3):304–9. doi:10.1016/j.ajem.2008.12.014.

6. Medley DB, Morris JE, Stone CK, et al. An association between occupancy rates in the emergency department and rates of violence toward staff. J Emerg Med. 2012;43(4):736–44. doi:10.1016/j.jemermed.2011.06.131.

7. Richardson DB. Increase in patient mortality at 10 days associated with emergency department overcrowding. Med J Aust. 2006;184(5):213–6. doi:10.5694/j.1326-5377.2006.tb00204.x.

8. Sharma A, Inder B. Impact of co-located general practitioner (GP) clinics and patient choice on duration of wait in the emergency department. Emerg Med J. 2011;28(8):658–61. doi:10.1136/emj.2009.086512.

9. Arnaud É, Elbattah M, Gignon M, et al. Deep learning to predict hospitalization at triage: integration of structured data and unstructured text. In: 2020 IEEE International Conference on Big Data (Big Data). IEEE; 2020. doi:10.1109/BigData50022.2020.9378073.

10. Sarasa Cabezuelo A. Application of machine learning techniques to analyze patient returns to the emergency department. J Pers Med. 2020;10(3):81. doi:10.3390/jpm10030081.

11. Ohno T. O sistema Toyota de produção: além da produção em larga escala. Porto Alegre: Bookman; 1997.

12. Mazzocato P, Stenfors-Hayes T, von Thiele Schwarz U, et al. Lean thinking in healthcare: a realist review of the literature. Qual Saf Health Care. 2010;19(5):376–82. doi:10.1136/qshc.2009.037986.

13. Isfahani HM, Tourani S, Seyedin H. Features and results of conducted studies using a lean management approach in emergency department in hospital: a systematic review. Bull Emerg Trauma. 2019;7(1):9. doi:10.29252/beat-070102.

14. Tlapa D, Zepeda-Lugo CA, Tortorella GL, et al. Effects of lean healthcare on patient flow: a systematic review. Value Health. 2020;23(2):260–73. doi:10.1016/j.jval.2019.11.002.

15. Souza DL, Korzenowski AL, Alvarado MM, et al. A systematic review on lean applications in emergency departments. Healthcare (Basel). 2021;9(6):763. doi:10.3390/healthcare9060763.

16. Wang J, Lv H, Chen M, et al. A systematic review of Lean implementation in hospitals: impact on efficiency, quality, cost, and satisfaction. Int J Health Policy Manag. 2025;14:8974. doi:10.34172/ijhpm.8974.

17. Vermeulen MJ, Stukel TA, Guttmann A, et al. Evaluation of an emergency department lean process improvement program to reduce length of stay. Ann Emerg Med. 2014;64(5):427–38. doi:10.1016/j.annemergmed.2014.06.007.

18. Des Jarlais DC, Lyles C, Crepaz N; Trend Group. Improving the reporting quality of nonrandomized evaluations of behavioral and public health interventions: the TREND statement. Am J Public Health. 2004;94(3):361–6. doi:10.2105/AJPH.94.3.361.

19. Brasil. Ministério da Saúde; Secretaria de Atenção Especializada à Saúde; CONASS; CONASEMS. Lean nas Emergências: guia prático para transformação e eficiência operacional na saúde. Brasília, DF: Ministério da Saúde; 2026.

20. Callaway B, Sant’Anna PHC. Difference-in-differences with multiple time periods. J Econom. 2021;225(2):200–30. doi:10.1016/j.jeconom.2020.12.001.

21. Chen M, Guan Q, Zhuang J. Patient-centered lean healthcare management from a humanistic perspective. BMC Health Serv Res. 2024;24(1):1261. doi:10.1186/s12913-024-11755-w.

22. Costa LBM, Godinho Filho M, Rentes AF, et al. Lean healthcare in developing countries: evidence from Brazilian hospitals. Int J Health Plann Manage. 2017;32(1):e99–e120. doi:10.1002/hpm.2331.

